# Public health interventions in India slowed the spread of COVID-19 epidemic dynamics

**DOI:** 10.1101/2020.06.06.20123893

**Authors:** Balbir B Singh, Mark Lowerison, Ryan T. Lewinson, Isabelle A. Vallerand, Rob Deardon, Játinder PS Gill, Baljit Singh Gill, Herman W Barkema

## Abstract

**Background:** The government of India implemented social distancing interventions to contain the COVID-19 epidemic. However, effects on epidemic dynamics are yet to be understood.

**Methods:** Rates of laboratory-confirmed COVID-19 infections per day and effective reproduction number (*R_t_*) were estimated for 4 periods (Pre-lockdown and Lockdown Phases 1 to 3) according to nationally implemented phased interventions. Adoption of these interventions was estimated using Google mobility data. Estimates at the national level and for 12 Indian states most affected by COVID-19 are presented.

**Findings:** Daily case rates ranged from 0·03 to 30·05/10 million people across 4 discrete periods in India. From May 4-17, 2020, the National Capital Territory (NCT) of Delhi had the highest case rate (222/10 million people/day), whereas Kerala had the lowest (2·18/10 million/day). Average *R_t_* was 1·99 (95% CI 1·93-2·06) for India; it ranged from 1·38 to 2·78, decreasing over time. Median mobility in India decreased in all contact domains, with the lowest being 21% in retail/recreation (95% CI 13-46%), except home which increased to 129% (95% CI 117-132%) compared to the 100% baseline value.

**Interpretation:** The Indian government imposed strict contact mitigation, followed by a phased relaxation, which slowed the spread of COVID-19 epidemic progression in India. The identified daily COVID-19 case rates and *R_t_* will aid national and state governments in formulating ongoing COVID-19 containment plans. Furthermore, these findings may inform COVID-19 public health policy in developing countries with similar settings to India.

**Funding:** Non-funded.

## Research in context

### Evidence before this study

The first imported case of COVID-19 was reported in India on 30 January 2020. Within < 4 mo (21 May 2020), the country had 112,359 cumulative cases and 3,435 cumulative deaths. The Indian government implemented several non-pharmaceutical interventions to contain the COVID-19 epidemic. However, adherence to these interventions and effects on epidemic dynamics remain unknown.

### Added value of this study

The rate of laboratory-confirmed COVID-19 infections continued to rise during lockdown Phases 1 to 3, despite the government-imposed interventions. However, the effective reproduction number (*R_t_*) decreased from 2·78 in the Lockdown Phase 1 to 1·38 in the Lockdown Phase 3. Using Google mobility data, we inform that mobility in India decreased (compared to the 100% baseline value) in all contact domains except home.

### Implications of all available evidence

Adoption level of the public health contact mitigation interventions imposed by the Indian government was higher than in many other countries. However, the persistence of *R_t_* above 1.0 during the lockdown Phases 1 to 3 indicated that moderate interventions will be unable to halt and eventually eradicate the COVID-19 epidemic from India.

## INTRODUCTION

Coronavirus disease 2019 (COVID-19) was declared a pandemic by the WHO on March 11, 2020^1^. This novel infection, caused by the severe acute respiratory syndrome coronavirus 2 (SARS-CoV-2), was first reported in Wuhan, China^2^ and is the seventh coronavirus known to infect humans^3^. As of 05 June 2020, 6,535,354 cases and 387,155 deaths have been reported worldwide^4^. As data collection efforts and testing protocols continue to evolve, these values likely represent a considerable underestimation of total cases and deaths^5^.

The successful impact of public health interventions on COVID-19, including social distancing, isolating infected individuals and quarantine in Wuhan, China, supported implementation of similar measures in many other countries^6^. For instance, mass gathering events have been reported to pose a considerable public health risk and therefore have been avoided in most COVID-19 infected countries. Based on modelling studies, whereas highly effective contact tracing coupled with case isolation has potential to contain COVID-19 outbreaks, proper containment likely requires additional measures, e.g. contact mitigation^7^. However, with recent declines in COVID-19 case volume, many countries are turning to contact mitigation relaxation plans.

The federal government in India responded swiftly to COVID-19 by implementing staged lockdown periods across the country; strict, intense contact mitigation was implemented initially, followed by phased relaxation as deemed appropriate. As India is among the world’s most populated countries, COVID-19 has considerable potential for widespread morbidity and mortality and containment has global implications. Evaluating impacts of COVID-19 public health interventions is important to inform future effective public health and social interventions. The effective reproduction number (*R_t_*), which captures time-dependent variations in the transmission potential of an infectious disease in a given population, is an important parameter for evaluating effectiveness of public health interventions^8^. Although epidemiological characteristics of COVID-19, including rates of confirmed cases and *R_t_*, were investigated in Wuhan, China across various periods of social distancing interventions^8^, the impact of government-imposed mitigation strategies on transmission dynamics in India remains unknown. In this study, we estimated the laboratory-confirmed daily case rate, and *R_t_* of COVID-19 epidemic in key lockdown periods in India. Estimates presented here will provide key information for ongoing COVID-19 prevention and control in India.

## METHODS

### Source of data

Informed consent for collection of epidemiological data was not required, as these data were already coded and available in the public domain. No identifiable personal information was used in this study.

#### Time-series data

Indian COVID-19 time series incidence and fatality data were extracted on May 24, 2020 using the WHO Coronavirus disease 2019 (COVID-19) situation reports 10-125^9^. We selected data from states/union territories where > 200 COVID-19 cases (positive/recovered/deceased) had been recorded before May 22, 2020, which included data from 17 states/union territories. Time-series data on daily counts were not available for 5 states (Gujarat, Rajasthan, Uttar Pradesh, Jammu and Kashmir, and Bihar); therefore, these were excluded from the study. Time-series data for the remaining 12 states were extracted from official state Health and Family Welfare department websites (Appendix P 2).

#### Census data

The official human population 2011 Indian census data were used^10^. In 2014, the state of Andhra Pradesh was bifurcated into two states, Telangana and residuary Andhra Pradesh. As separate census data for these states were not available, combined results for Andhra Pradesh (Telangana and residuary Andhra Pradesh) are presented.

### Case definitions

Cases were defined based on laboratory confirmation using throat/nasal swab real-time reverse transcriptase–polymerase chain reaction (RT-PCR) assay, the diagnostic test recommended by the Indian Council for Medical Research (ICMR) for diagnosis of COVID-19^11^. Only laboratory-confirmed cases were included in the analyses. The ICMR strategy for COVID-19 testing included all symptomatic individuals who had undertaken international travel in the last 14 d, all symptomatic contacts of laboratory conformed cases, all symptomatic health care workers, all patients with severe acute respiratory illness (fever and cough and/or shortness of breath), and asymptomatic direct and high-risk contacts of a confirmed cases^11^. For hotspots/clusters, large migration gatherings and evacuation centres, all persons having fever, cough, sore throat, or runny nose were recommended to be tested^11^.

### Classification of 4 time periods

The study period comprised an initial unmitigated social contact period when temperature screening was imposed at international checkpoints. This was followed by strict contact mitigation and then phased relaxation. Dynamics of the COVID–19 epidemic were evaluated in discrete periods, according to changes in government policies (Fig. 1). Based on these government-imposed interventions, we classified 4 periods for analysis, including: 1) Pre-lockdown; Unmitigated social contact from January 30, 2020 (first reported case) to March 21, 2020 during which no COVID-19 specific interventions were imposed, and both imported, and locally transmitted cases were reported; 2) Lockdown Phase 1; March 22, 2020 (day on which a 14-h voluntary public curfew was imposed) to April 14, 2020 (including a 21-d nationwide lockdown from March 24, 2020 to April 14, 2020); 3) Lockdown Phase 2; April 15, 2020 to May 03, 2020 (19-d nationwide lockdown); and 4) Lockdown Phase 3; May 4, 2020 to May 17, 2020 (14-d nationwide lockdown). Detailed restrictions during nationwide Lockdowns Phases 1 to 3 are available at the website of the Ministry of Home Affairs, Government of India (https://www.mha.gov.in/media/whats-new, accessed on May 17, 2020). Briefly, conditional relaxation was allowed in Lockdown Phase 2 after April 20, 2020 in areas where spread was contained. During Lockdown Phase 3, the Indian government divided the country into green (districts with no confirmed case in last 21 d), orange (districts that were in neither red nor green zones) and red zones (hotspot districts), with relaxations accordingly.

**Fig. 1.**
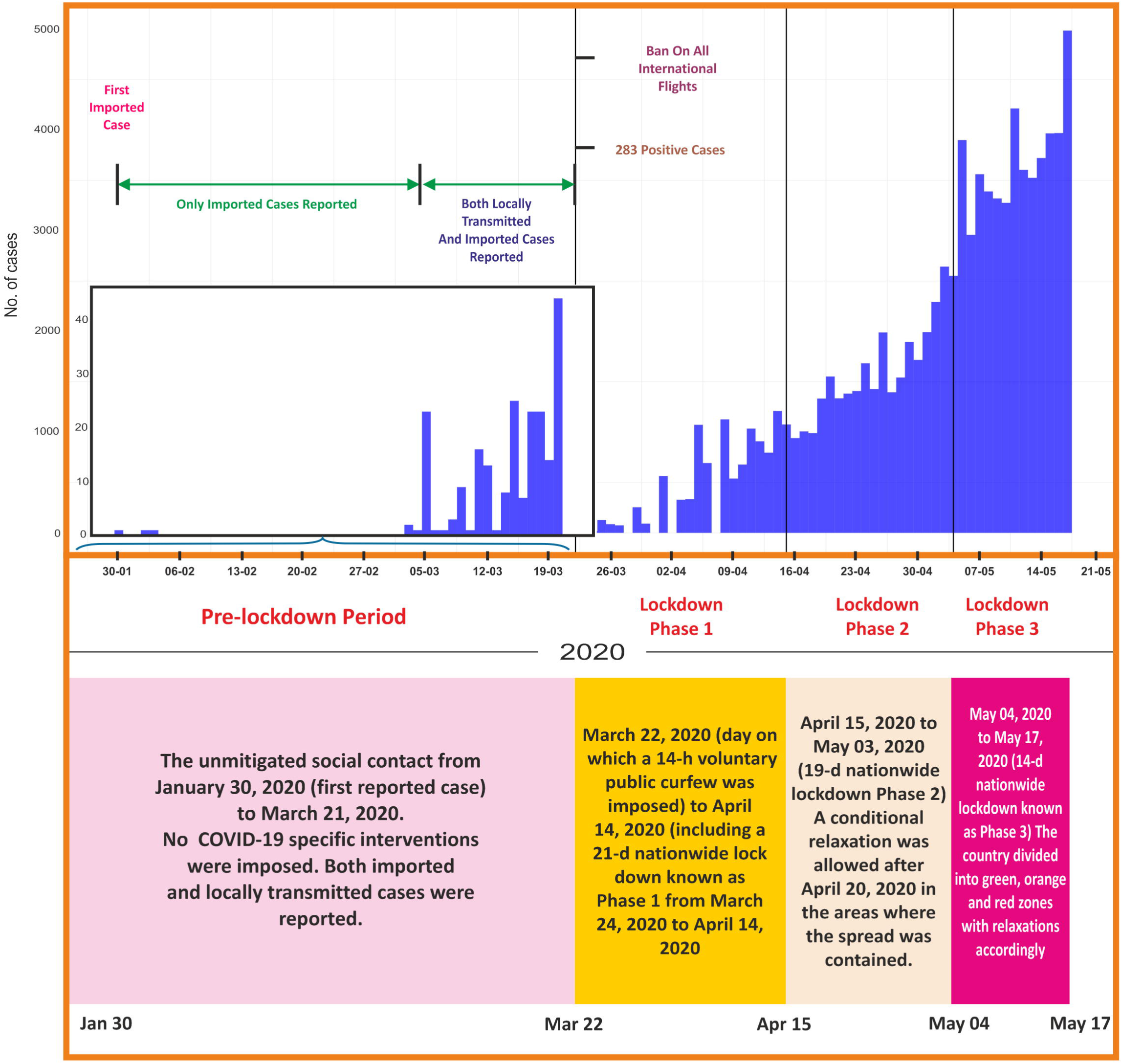
Incidence of COVID–19 cases, key events and public health interventions across 4 periods in India. Detailed information relating to restrictions and relaxations during the nationwide lockdowns Phase 1 to 3 are available at the website of the Ministry of Home Affairs, Government of India (https://www.mha.gov.in/media/whats-new).

### Mobility index

A domain-specific mobility index was constructed using India’s mobility report (Google Inc., Mountain View, CA, USA)^12^. These data are publicly available from Google and represent the percent change from baseline mobility within various domains (retail and recreation mobility, grocery and pharmacy mobility, parks mobility, transit stations mobility, workplace mobility, and residential mobility) according to cell phone-user geolocation data. As data were available as percent mobility change from the baseline value, we considered 100 as the baseline value; therefore, 100 was added to each value to transform the raw Google data to domain-specific mobility per day. The domain-specific mobility index was constructed for the country and 12 Indian states.

### Outcomes

Daily rate of laboratory-confirmed cases per 10 million people per day was estimated across periods for the 12 Indian states and for the country. We used number of cases in each period, divided by the number of days in each period (52, 24, 19 and 14 d) and the total population of the selected region as per the 2011 census^10^. The *R_t_* was calculated to determine the transmission of SARS-CoV-2 in each of the 4 periods. The *R_t_* was calculated based on the method^13^ that is able to detect changes in the *R_t_* following public health interventions.

### Statistical analyses

All statistical analyses were conducted using R version 3·6·3 (R Development Core Team, http://www.r-project.org). Epidemic curves were plotted, based on laboratory diagnosis date and intervention periods described. Choropleth maps describing geographical distributions of COVID–19 case rates for the country and 12 Indian states through 4 intervention periods were generated by R software.

The *R_t_* for 12 Indian states and for the entire country were calculated as per the method developed by Cori et al.^13^. This method estimates *R_t_* from the incidence time-series and incorporates uncertainty in the distribution of the serial interval. We used the daily number of reported COVID–19 cases from the above-mentioned official data sources. The serial interval required to estimate *R_t_* (mean = 7·5 d, SD = 3·4 d) was derived from previous studies in Wuhan, China^8,14^. The serial interval was considered constant across all periods. A 5-d moving average was used to estimate *R_t_* and its 95% credible interval on each day.

The first locally transmitted COVID-19 case is thought to have occurred in India on March 5, 2020 and the Indian government closed international borders and air travel on March 22, 2020; therefore, March 22 was taken as our first day of *R_t_* estimation^19^. As per Cori et al., this allows for a complete serial interval from the first locally transmitted case, and enables presumption of a closed population and ensures an appropriate total case count^13^. Therefore, the *R_t_* was estimated from March 22 to May 24 and results of initial burn-in period (when both imported and locally transmitted cases were reported; Fig. 1) are not presented. For the 12 states, *R_t_* was estimated for the whole period, but was presented from the day when 50 cumulative cases were reported (or from March 22 onwards, whichever is later) given the limited diagnostic cases and capacity in the preliminary period. Although country and state populations were assumed to be closed from March 22 onwards, the possibility of imported cases being reported cannot be ruled out at the state level.

Descriptive analyses to report changes in mobility index were conducted according to key intervention periods.

## RESULTS

### Epidemic curve

The COVID-19 epidemic started in India on January 30, 2020 with the first imported case detected^15^. Within < 4 mo, the country reported 5,600 new cases per day, 112,359 cumulative cases and 3,435 cumulative deaths on 21 May 2020^4^. COVID-19 has been reported from 33 of the 36 states/union territories of the country^16^.

Number of cases were minimal during the Pre-Lockdown Period but continued to rise in subsequent Lockdown periods (Phases 1-3), with approximately 5,000 cases/day reported at the end of Lockdown Phase 3 (Fig. 1). There were obvious differences among states in epidemic progression (Fig. 2).

**Fig. 2.**
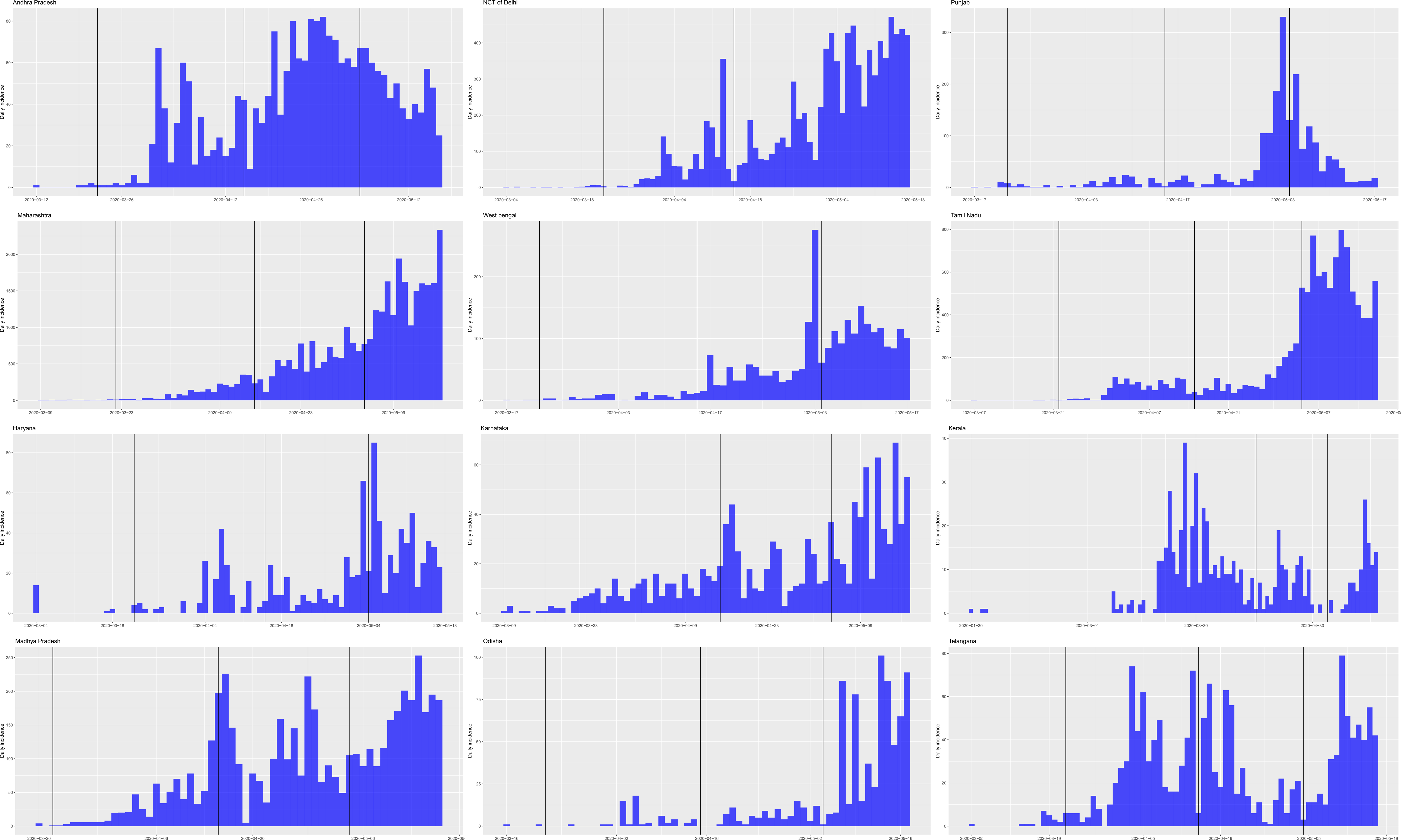
Incidence of COVID–19 cases across 4 periods in 12 states of India. Vertical bars illustrate 4 periods related to public health interventions in India. Discrete periods: 1) Pre-lockdown - January 30, 2020 to March 21, 2020; 2) Lockdown Phase 1 - March 22, 2020 to April 14, 2020; 3) Lockdown Phase 2 - April 15, 2020 to May 3, 2020 and 4) Lockdown Phase 3 - May 4, 2020 to May 17, 2020.

As of 17 May 2020, the state of Maharashtra had the highest number of cumulative cases (n=33,043) followed by Tamil Nadu (n=11,001) and NCT of Delhi (n=9751). Kerala reported the highest number of cases per day during Lockdown Phase 1. The highest number of cases/day were reported in Andhra Pradesh, Punjab and West Bengal during Lockdown Phase 2. Over 1,500 cases/d were reported from Maharashtra at the end of Lockdown Phase 3, followed by NCT of Delhi and Tamil Nadu where close to 400 cases/d were reported (Fig. 2).

### Case rate/10 million people/day

The case rate was 0·03, 3·50, 12·87, and 30·05 per 10 million people per day across the 4 discrete periods in India. Overall, the case rate was 7 per 10 million people per day from 30 January 2020 to 17 May 2020. The rate of increase was maximal from Pre-lockdown Period through Lockdown Phase 1 and minimal from Lockdown Phase 2 through Phase 3 in the country (Table 1).

**Table 1.**
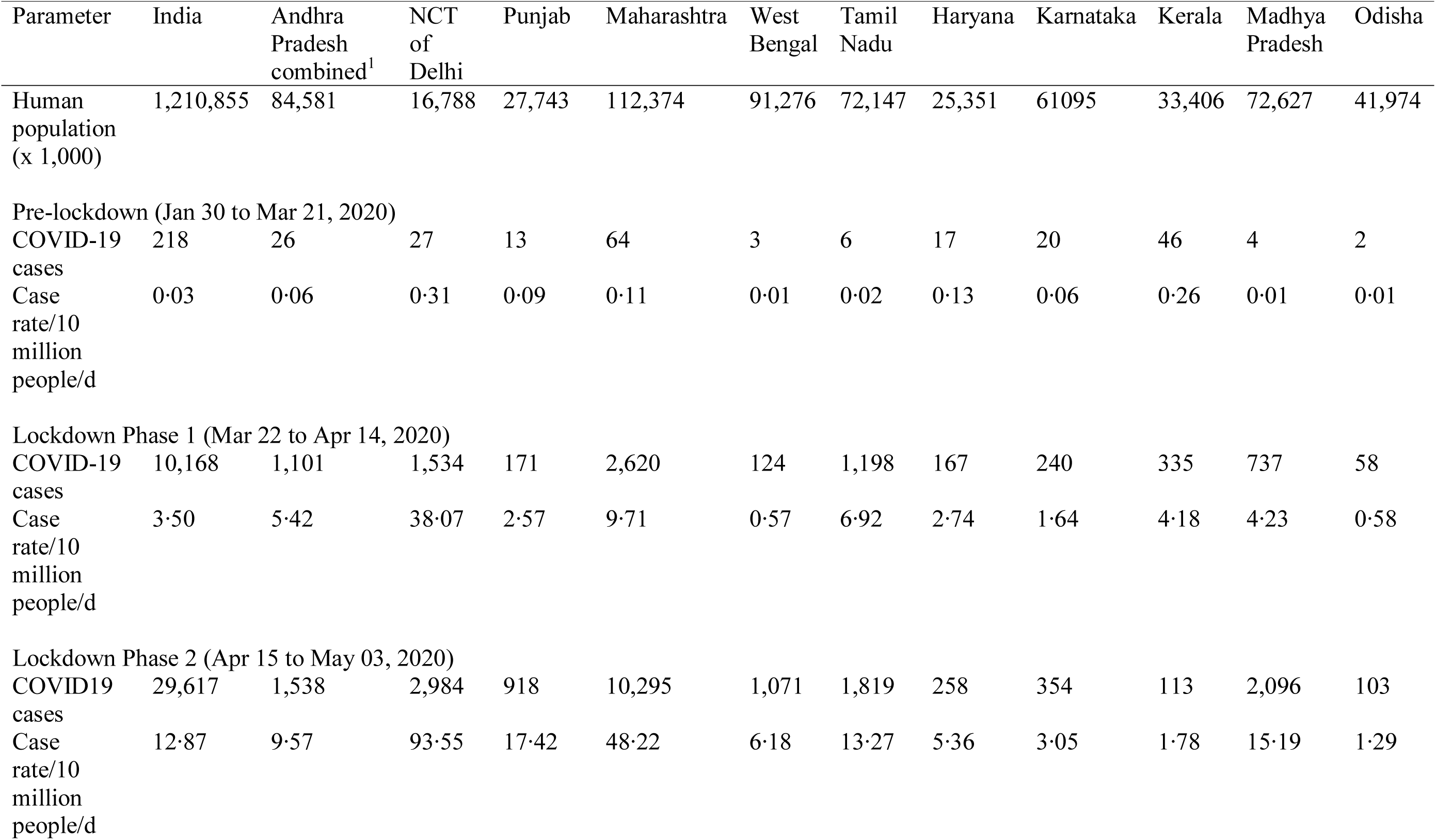
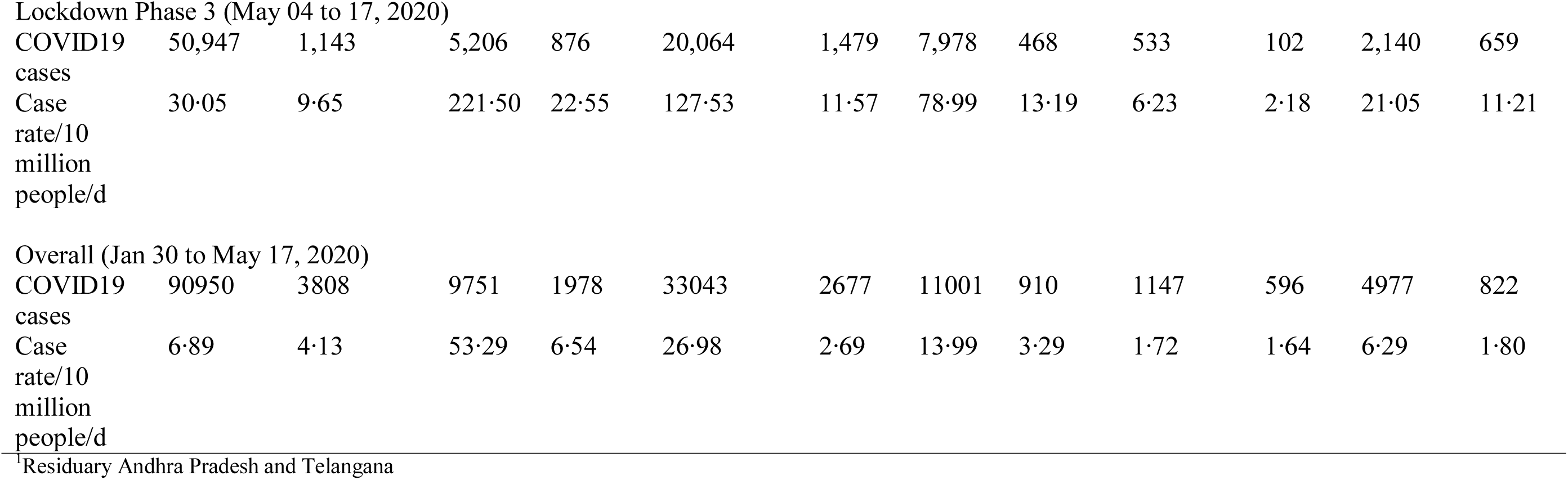
Daily incidence of COVID-19 cases during the 4 Periods in 12 states of India.

| Parameter | India | Andhra Pradesh combined<sup>1</sup> | NCT of Delhi | Punjab | Maharashtra | West Bengal | Tamil Nadu | Haryana | Karnataka | Kerala | Madhya Pradesh | Odisha |
| --- | --- | --- | --- | --- | --- | --- | --- | --- | --- | --- | --- | --- |
| Human population (x 1,000) | 1,210,855 | 84,581 | 16,788 | 27,743 | 112,374 | 91,276 | 72,147 | 25,351 | 61,095 | 33,406 | 72,627 | 41,974 |
| Pre-lockdown (Jan 30 to Mar 21, 2020) |  |  |  |  |  |  |  |  |  |  |  |  |
| COVID-19 cases | 218 | 26 | 27 | 13 | 64 | 3 | 6 | 17 | 20 | 46 | 4 | 2 |
| Case rate/10 million people/d | 0.03 | 0.06 | 0.31 | 0.09 | 0.11 | 0.01 | 0.02 | 0.13 | 0.06 | 0.26 | 0.01 | 0.01 |
| Lockdown Phase 1 (Mar 22 to Apr 14, 2020) |  |  |  |  |  |  |  |  |  |  |  |  |
| COVID-19 cases | 10,168 | 1,101 | 1,534 | 171 | 2,620 | 124 | 1,198 | 167 | 240 | 335 | 737 | 58 |
| Case rate/10 million people/d | 3.50 | 5.42 | 38.07 | 2.57 | 9.71 | 0.57 | 6.92 | 2.74 | 1.64 | 4.18 | 4.23 | 0.58 |
| Lockdown Phase 2 (Apr 15 to May 03, 2020) |  |  |  |  |  |  |  |  |  |  |  |  |
| COVID19 cases | 29,617 | 1,538 | 2,984 | 918 | 10,295 | 1,071 | 1,819 | 258 | 354 | 113 | 2,096 | 103 |
| Case rate/10 million people/d | 12.87 | 9.57 | 93.55 | 17.42 | 48.22 | 6.18 | 13.27 | 5.36 | 3.05 | 1.78 | 15.19 | 1.29 |
| Lockdown Phase 3 (May 04 to 17, 2020) |  |  |  |  |  |  |  |  |  |  |  |  |
| COVID19 cases | 50,947 | 1,143 | 5,206 | 876 | 20,064 | 1,479 | 7,978 | 468 | 533 | 102 | 2,140 | 659 |
| Case rate/10 million people/d | 30.05 | 9.65 | 221.50 | 22.55 | 127.53 | 11.57 | 78.99 | 13.19 | 6.23 | 2.18 | 21.05 | 11.21 |
| Overall (Jan 30 to May 17, 2020) |  |  |  |  |  |  |  |  |  |  |  |  |
| COVID19 cases | 90950 | 3808 | 9751 | 1978 | 33043 | 2677 | 11001 | 910 | 1147 | 596 | 4977 | 822 |
| Case rate/10 million people/d | 6.89 | 4.13 | 53.29 | 6.54 | 26.98 | 2.69 | 13.99 | 3.29 | 1.72 | 1.64 | 6.29 | 1.80 |

There were large differences in incidence rates across 12 Indian states and across time periods (Table 1; Fig. 3). Among all states, the highest case rate was 53 per 10 million people per day from NCT of Delhi, whereas the lowest case rate was 2 per 10 million people per day from Kerala from 30 January 2020 to 17 May 2020 (Table 1).

**Figs. 3.**
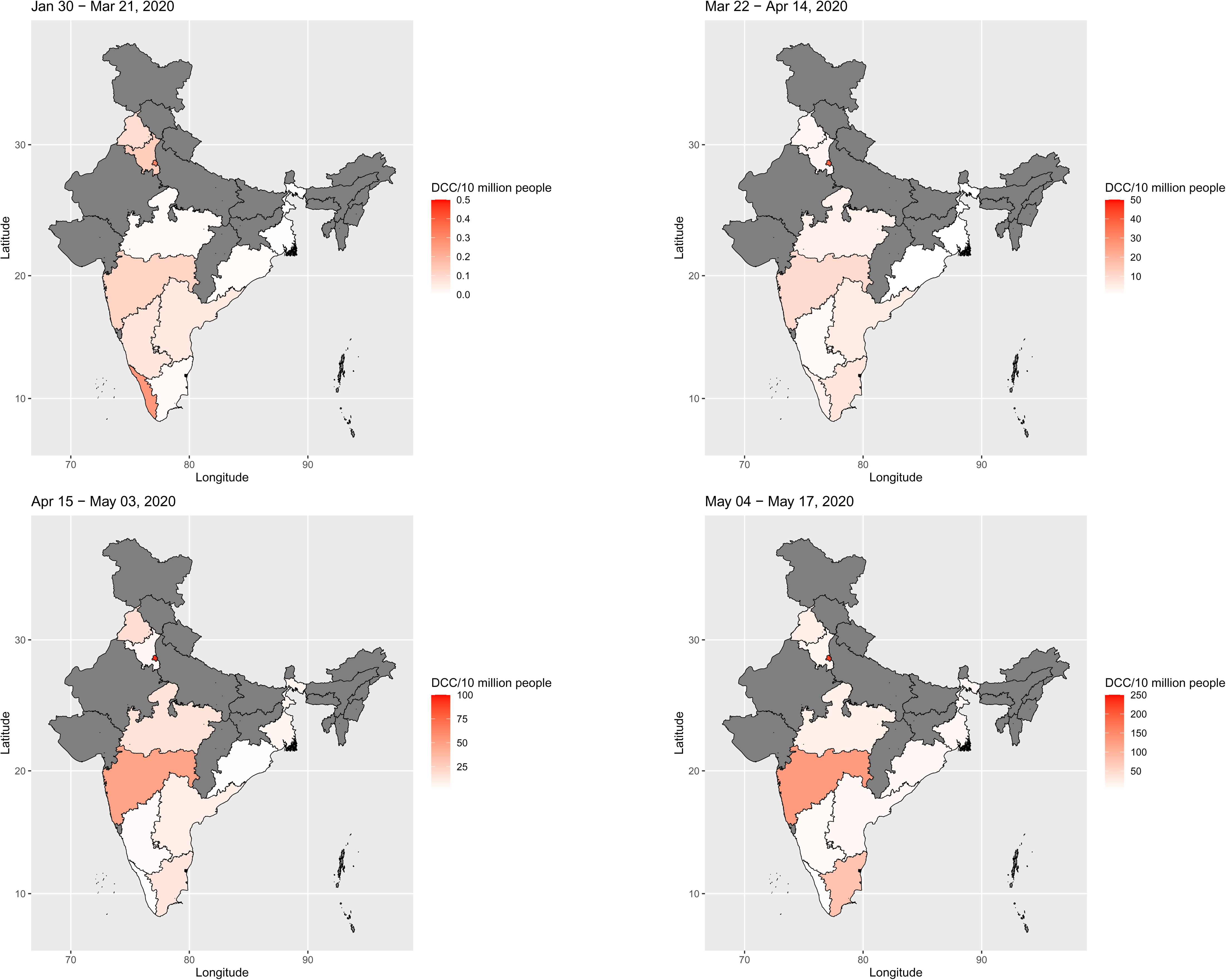
Geographic distribution of incidence of COVID-19 cases during the 4 Periods in 12 states of India. Note that combined results of Andhra Pradesh (for 2 states residuary Andhra Pradesh and Telangana) have been presented. DCC represents Daily Confirmed Cases, expressed as number of laboratory-confirmed cases per 10 million people per day.

In Lockdown Phase 3, the highest case rate was recorded for NCT of Delhi (222 per 10 million people per day) and the lowest case rate was recorded for Kerala (2·18 per 10 million people per day). The case rate for all states continued to rise despite implementing public health interventions, except Kerala, where case rate decreased from Lockdown Phase 1 to Phase 3 (Fig. 3).

### Effective Reproduction number

The *R_t_* differed across all 4 periods. Average *R_t_* was 1·99 (95% CI 1·93–2·06) for the entirety of India during the period from 22 March 2020 to 17 May 2020. The *R_t_* was 2·78 (95% CI 2·65–2·91) in Lockdown Phase 1, 1·45 (95% 1·42–1·47) in Phase 2, and 1·38 (95% CI 1·36–1·40) in Phase 3. Therefore, the *R_t_* had a consistent decreasing trend in India from Periods 2 through 4, respectively (Fig. 4, Appendix p 3–4).

**Fig. 4.**
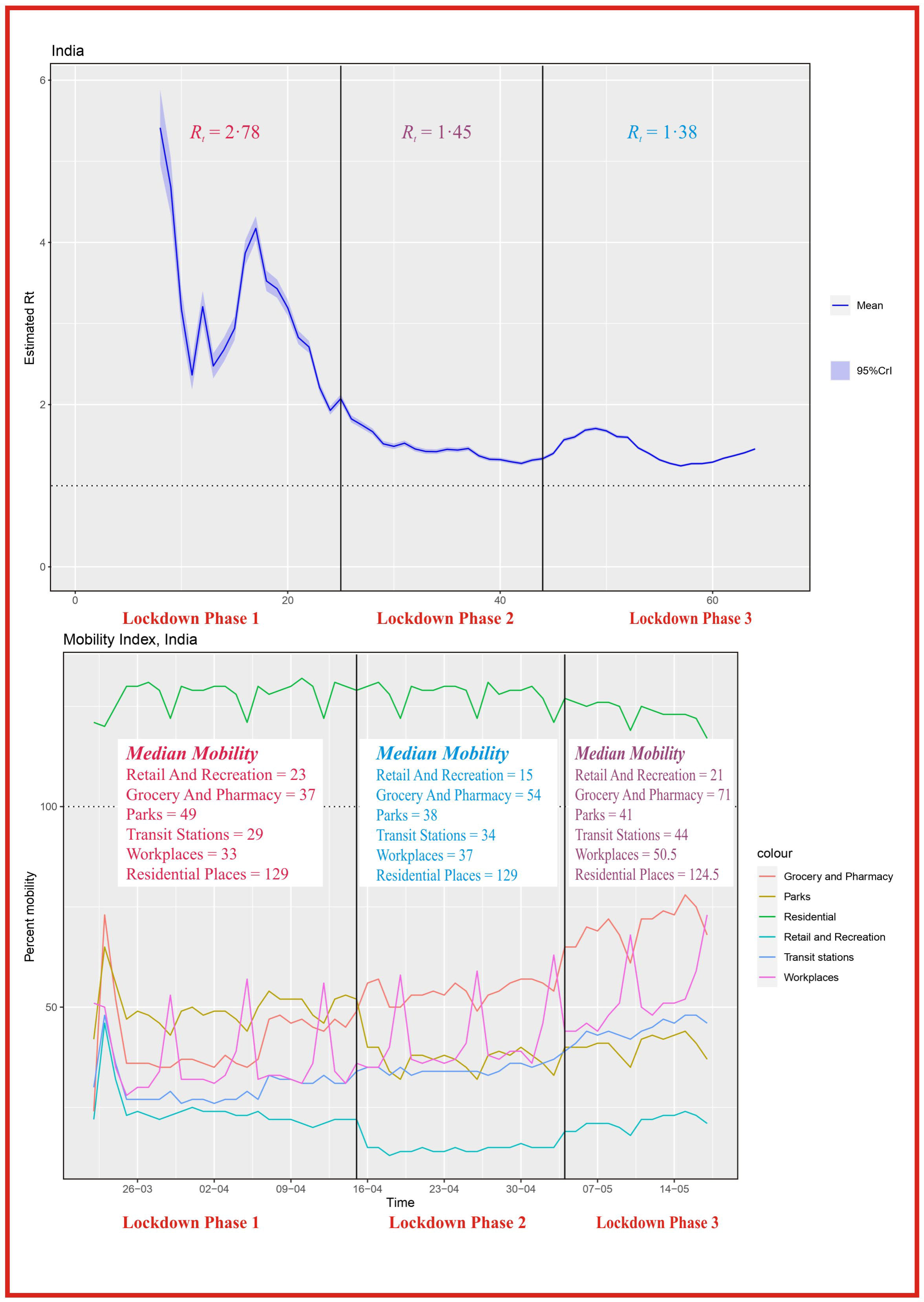
The effective reproduction number (*R_t_*) for the laboratory-confirmed COVID-19 cases and the domain-specific mobility of Indian citizens across 3 discrete periods (Lockdown Phases 1-3) in India. Vertical bars illustrate 3 discrete periods related to public health interventions in India. Discrete periods: 2) Lockdown Phase 1 - March 22, 2020 to April 14, 2020; 3) Lockdown Phase 2 - April 15, 2020 to May 3, 2020 and 4) Lockdown Phase 3 - May 4, 2020 to May 17, 2020. Note that 100 was considered as a baseline mobility value when no interventions were imposed.

Strong geographic differences were identified in *R_t_* across discrete periods in the country. For example, *R_t_* was > 2 for Maharashtra (2·16), Odisha (2·3), and Punjab state of India (2·15) from 30 January 2020 to 17 May 2020. The *R_t_* was > 1·5 for NCT of Delhi (1·92), Karnataka (1·58), Madhya Pradesh (1·81), Tamil Nadu (1·91), Telangana (1·91), and West Bengal (1·92) state from 30 January 2020 to 17 May 2020. The *R_t_* was < 1·5 for Kerala (1·48), Haryana (1·48), and Andhra Pradesh (1·27) from 30 January 2020 to 17 May 2020 (Fig. 5; Appendix 5–22).

**Fig. 5.**
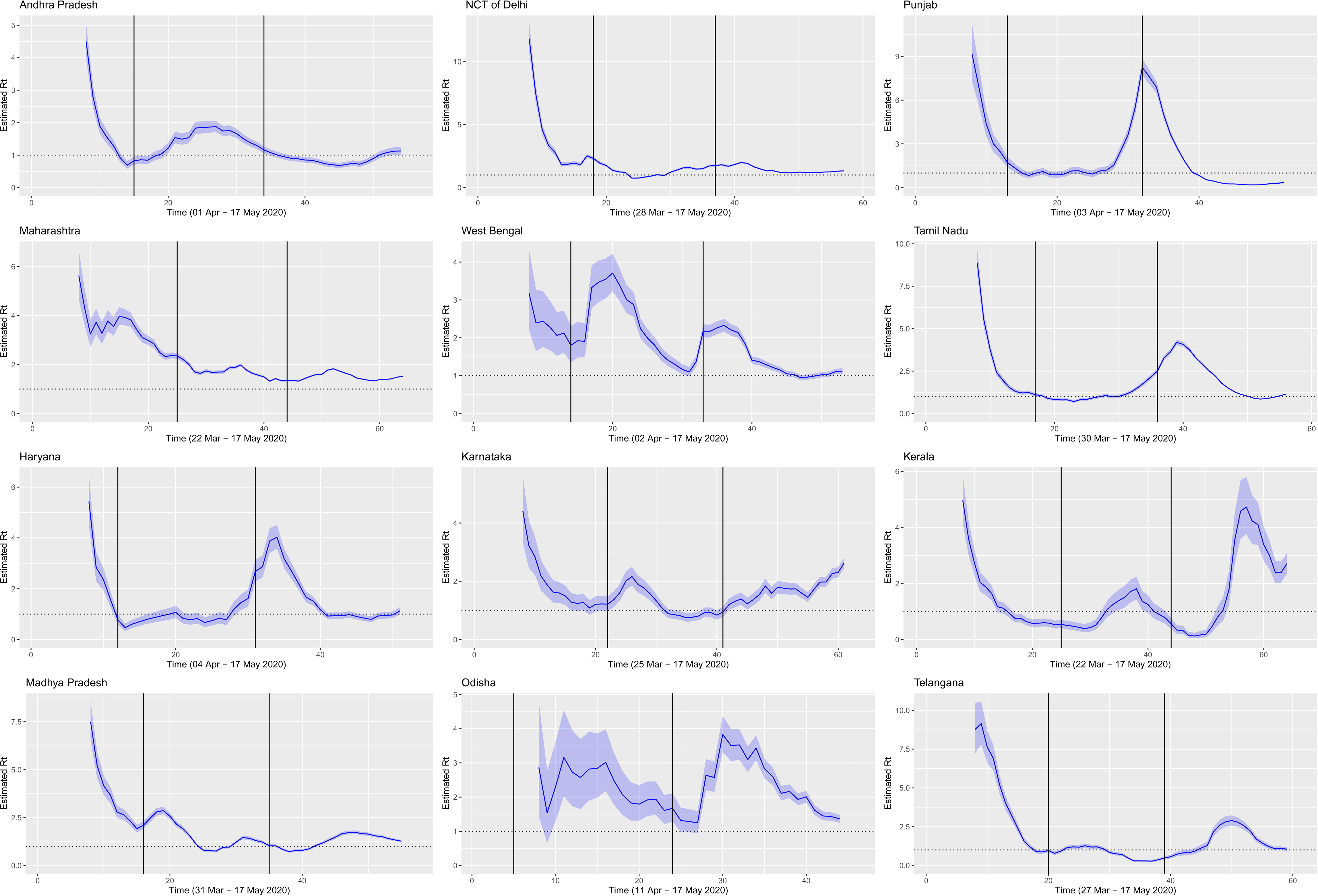
The effective reproduction number (*R_t_*) for laboratory-confirmed COVID-19 cases across 3 discrete periods (Periods 2-4) in 12 states of India. Vertical bars illustrate 3 discrete periods related to public health interventions in India. Discrete periods: 2) Lockdown Phase 1 - March 22, 2020 to April 14, 2020; 3) Lockdown Phase 2 - April 15, 2020 to May 3, 2020 and 4) Lockdown Phase 3 - May 4, 2020 to May 17, 2020. Note that starting point of the estimation varied in states subject to 50 cumulative cases reported. Overall, most states experienced a consistently decreasing trend from Lockdown Phases 1 through 3, respectively.

### Mobility index

Median mobility was 21% for retail and recreation, 53% at the grocery and pharmacy, 42% at parks, 34% at transit stations, 38% at workplaces and 129% at residential places from 22 March 2020 to 17 May 2020 (Table 2). Mobility at grocery and pharmacy (median 37%), transit stations (median 29%) and workplaces (median 33%) was lowest from 22 March 2020 to 14 April 2020, during Lockdown Phase 1 (Fig. 4). Interestingly, mobility for retail and recreation (median 15%) and Parks (median 38%) was lowest from 15 April 2020 to 3 May 2020, during Lockdown Phase 2 (representing relaxation). Note that these values were derived in comparison to a 100% baseline mobility in the country/state value when no such interventions were imposed.

**Table 2.**
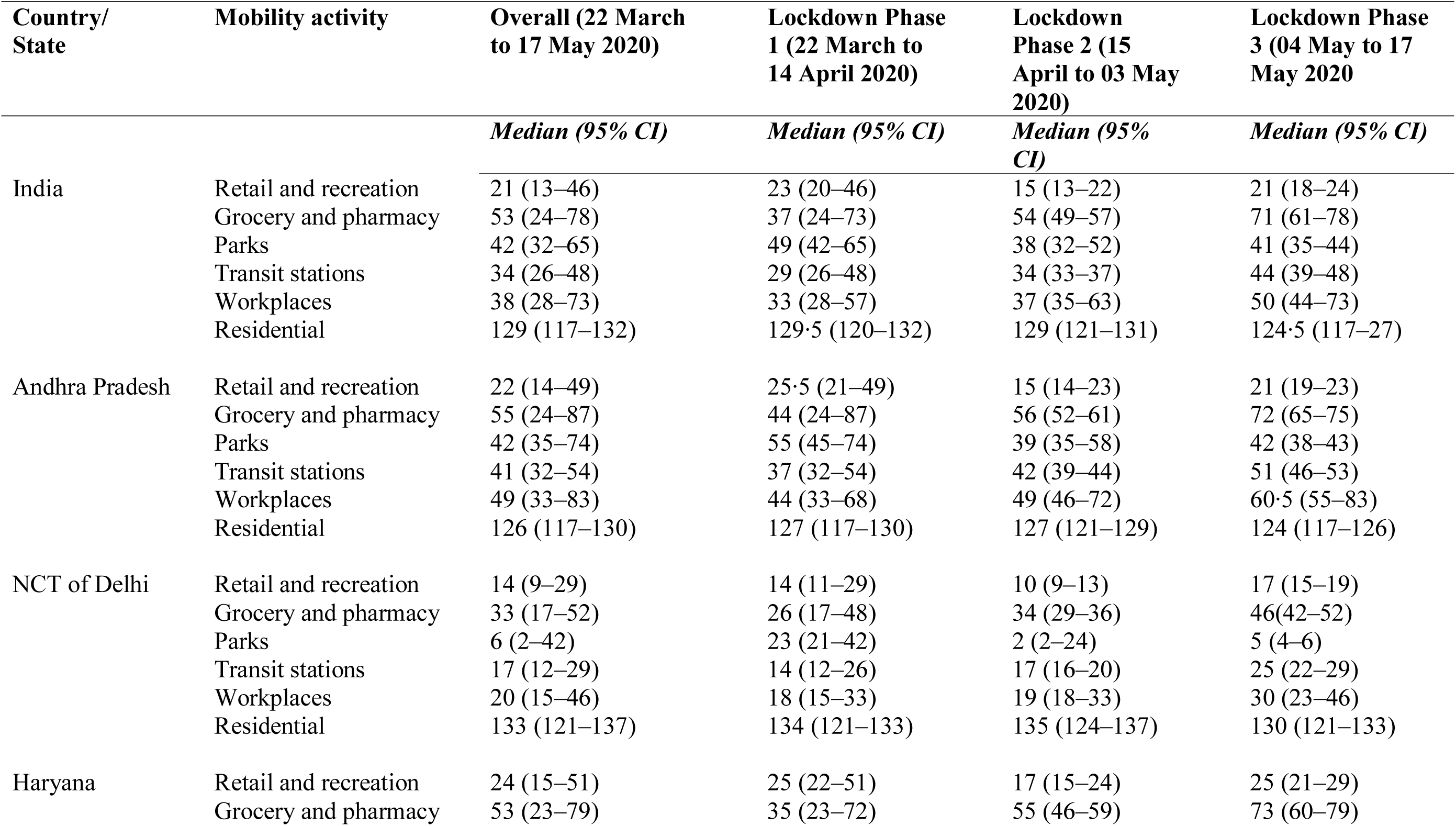
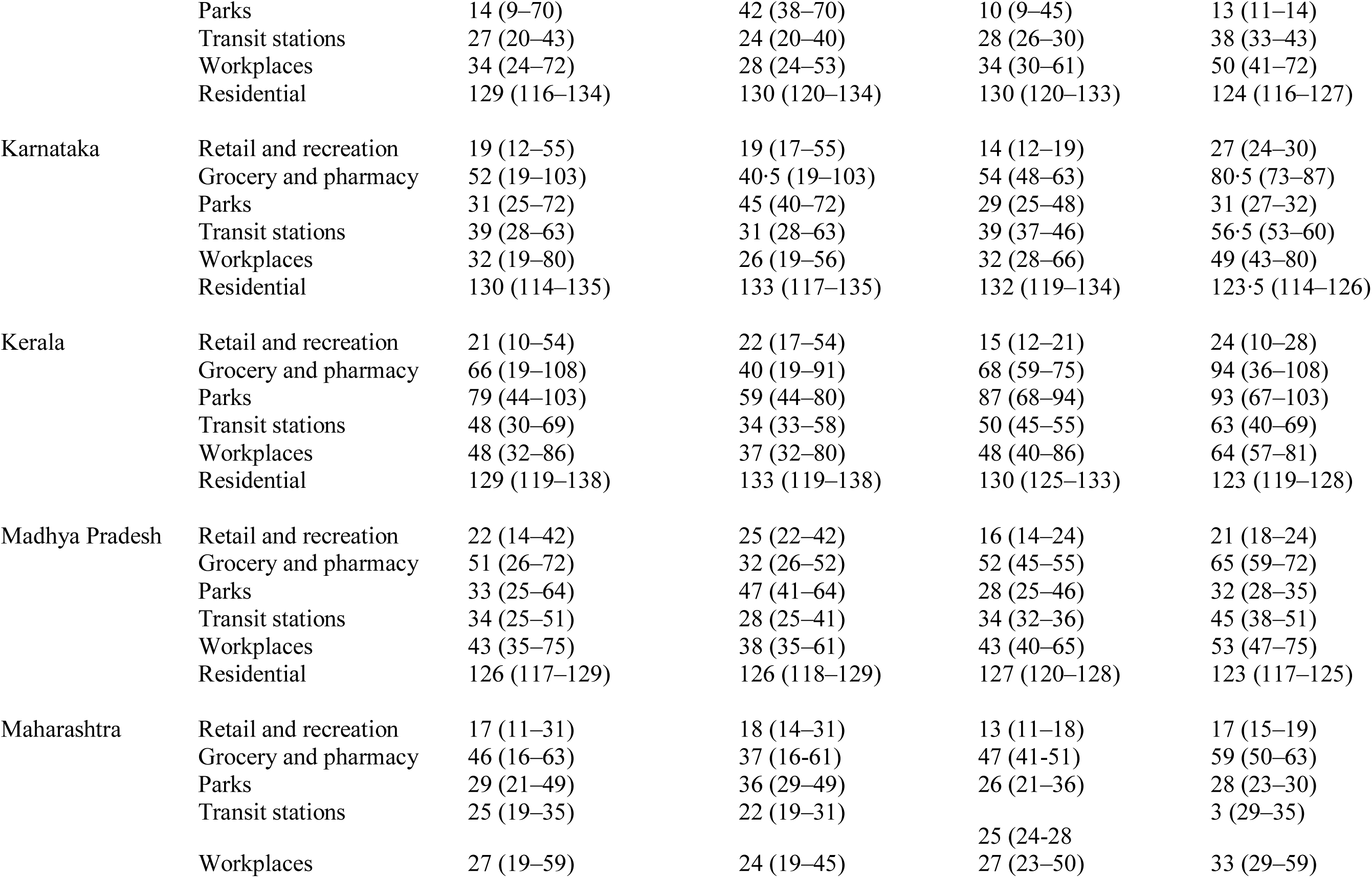
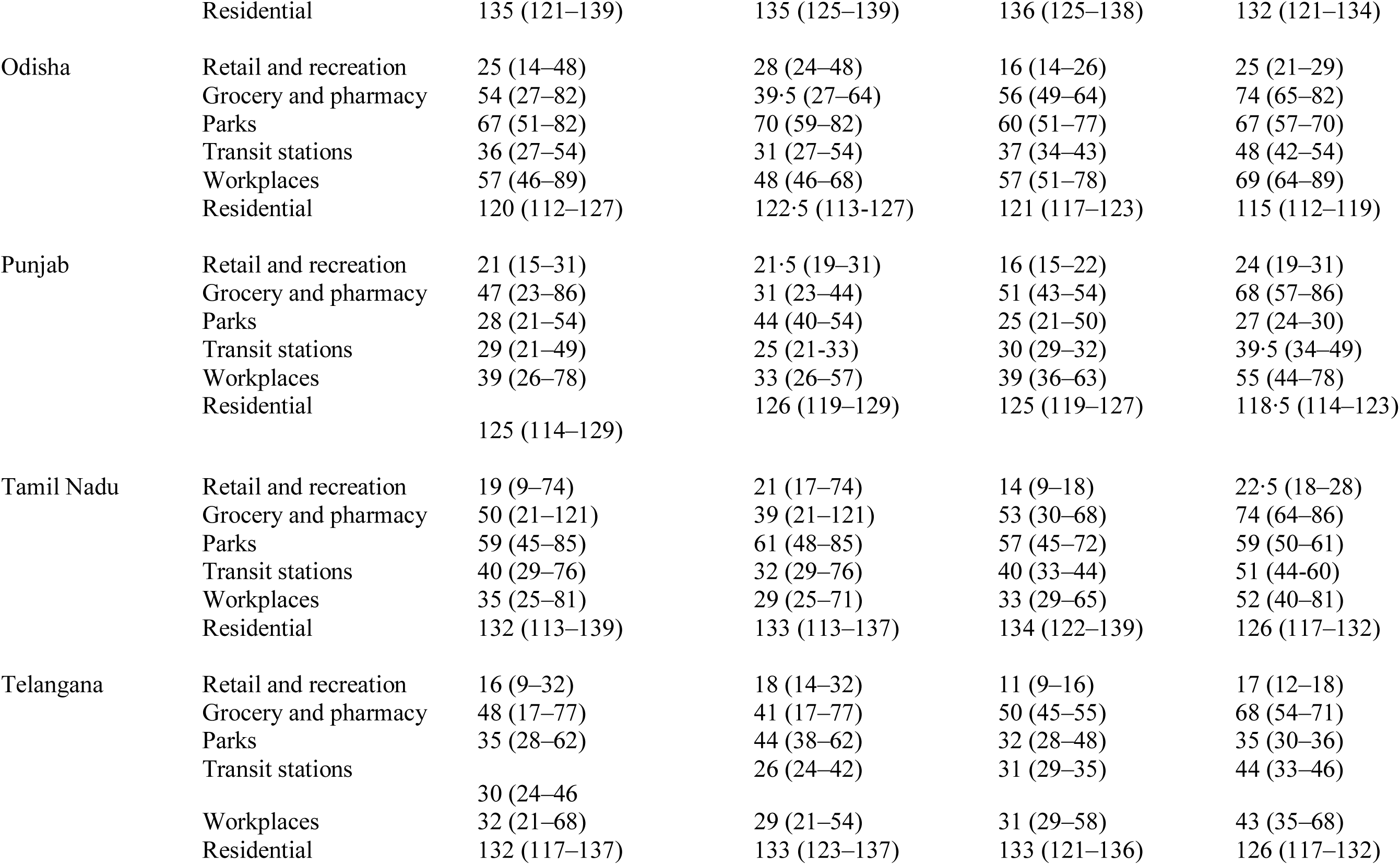
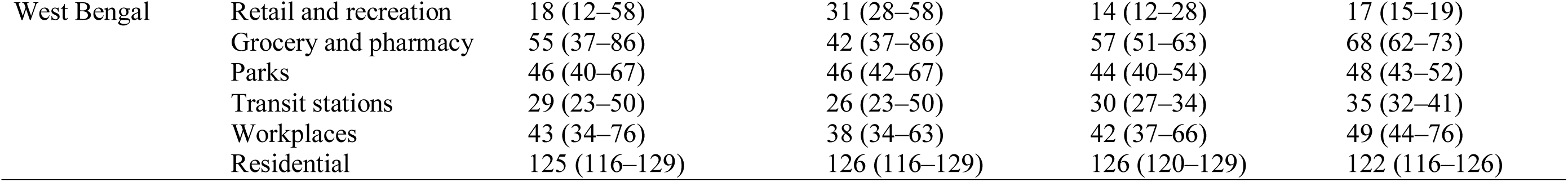
Domain-specific mobility in the country and 12 Indian states during key intervention periods. Note that a 100% baseline mobility was considered when no interventions were imposed.

The NCT of Delhi had the lowest mobility among the 12 states when compared to 100% baseline value at the retail and recreation (median 14%), grocery and pharmacy (median 33%), parks (median 6%), transit stations (median 17%), and workplaces (median 20%) from 22 March 2020 to 17 May 2020 (Fig. 6; Table 2).

**Fig. 6.**
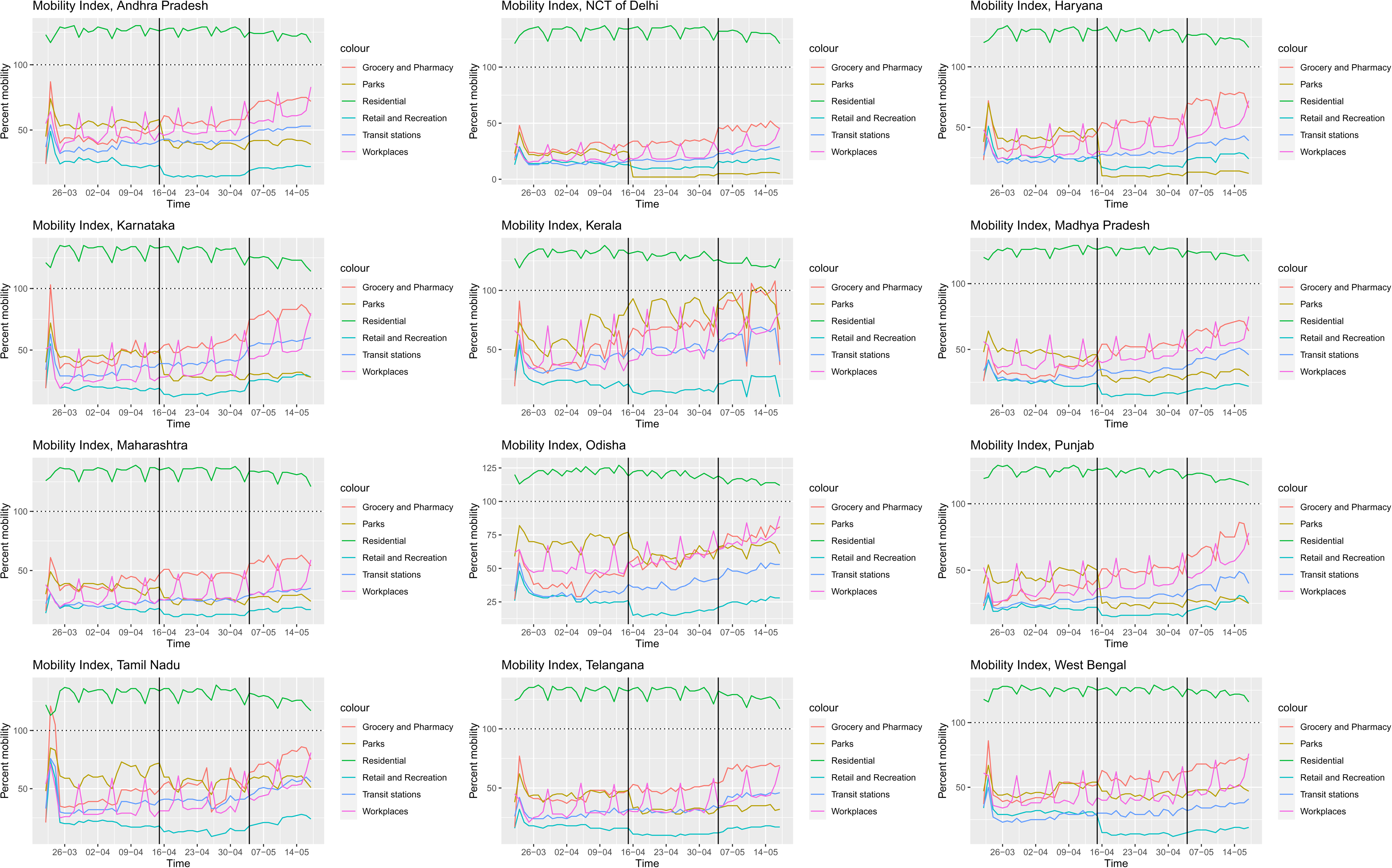
Domain-specific mobility of Indian citizens across 3 discrete periods (Periods 2-4) in 12 states of India. Vertical bars illustrate 3 discrete periods related to public health interventions in India. Discrete periods: 2) Lockdown Phase 1 - March 22, 2020 to April 14, 2020; 3) Lockdown Phase 2 - April 15, 2020 to May 3, 2020 and 4) Lockdown Phase 3 - May 4, 2020 to May 17, 2020. Note that 100 was considered as a baseline mobility value when no interventions were imposed.

## DISCUSSION

The daily confirmed case rate increased substantially from 0·03 to 30·05 per 10 million people per day during the 4 discrete periods in India. Among the 12 states, the highest case rate was recorded for the NCT of Delhi (53 per 10 million people per day), whereas the lowest was recorded for Kerala (1·64 per 10 million people per day) from 30 January 2020 to 17 May 2020. Many factors, including population structure and differences in the social contact rate/person/day likely influenced progression of COVID-19 in the country. Key factors influencing transmission are population density and possibility to implement physical distancing. The NCT of Delhi has a high population density (11,297 persons/km^2^), with 97·5% of the total population residing in urban areas and 10·91% of the urban population residing in slum areas^10^. In contrast, Kerala has a population density of 860 persons/km^2^, and 47·7% of the total population resides in urban areas and only 1·27% of the urban population resides in slum areas^10^. Social contact rate per person per day varied substantially in these settings in India: 17.0 in rural^17^, 28·3 in urban developed centres and 67·4 in urban slum areas^18^.

The case rate was lowest in Kerala state; it successfully implemented various measures, including testing, contact tracing, medical resource mobilisation and communication in the initial stage of the epidemic entry^19^. Additionally, Kerala had a Nipah virus outbreak during May 2018^20^ and gained experience in containment infectious disease outbreaks as compared to their counterpart states; this likely contributed to their low case rate.

The *R_t_* decreased in conjunction with implementation of public health measures including social distancing, travel restrictions, bans on mass gathering events, quarantining of positive cases and their contacts, and improved medical care. In Wuhan, China, the *R_t_* decreased from 3.0 in 26 January 2020 to 0·3 in 1 March 2020, within 40 d after multifaceted public health interventions were implemented^8^. For the United States, the *R_t_* reduced from 4·02 to 1·51 between March 17 and April 1, 2020^21^. As of 9 March 2020, the *R_t_* was reported to be 3·10 for Italy, 6·56 for France, 4·43 for Germany and 3·95 for Spain^22^. The reduction pattern in *R_t_* in India was similar to other areas of the world after implementing intense mitigation strategies. The *R_t_* in India was lower than in many European countries as well as the United States. As multiple factors were expected to have contributed to this change, associations of public health interventions with daily case rates and declining *R_t_* warrant further investigation.

Previous studies reported a varied basic reproduction number (*R_0_*) 1·03 – 4·18 in India^23–25^ due to differences in time periods, data sources or methods employed. The time-period in these studies was 4 March 2020 to 03 April 2020, with an estimated R_0_ of 2·56^24^, using initial epidemic growth phase data and reporting R_0_ to be 4·18^25^, and before 1 April 2020 and reporting R_0_ of 1·03. We used official data from 22 March 2020 to 17 May 2020 to derive *R_t_* estimates, so these estimates could not be compared.

The mobility index highlighted the adoption level of the public health contact mitigation interventions imposed by the Indian government; clearly, they were highly effective in substantially reducing social contact in the country. Mobility in the country was < 50% in all the domains except grocery and pharmacy, where mobility was 53% (as compared to 100% baseline value) from 22 March 2020 to 17 May 2020. This likely had an important role to contain COVID-19 epidemic in the country and will perhaps explain in part why our calculated *R_t_* for India was substantially lower than many European countries and the United States where mobility did not decrease as drastically as in India.

The current study had some limitations. Overall testing rate has not been very high in India; as of 23 May 2020, the country had tested 2,943,421 samples at a rate of 2,432 per million people. As of 1 June 2020, the United States had tested 17,612,125 samples at a rate of 53,911 per million people^26^. Information on the diagnostic testing patterns remain unknown. However, there was a shortage of testing during early phase of the epidemic. India tested 100 samples per day before 20 March 2020 but this was scaled-up to 100,000 tests per day on 20 May 2020^27^. It has been demonstrated that changes in testing rates affect the epidemic curve of COVID-19^28^. Therefore, an underestimated case rate in the initial stage of the epidemic cannot be ruled out. Additionally, migration of COVID-19 cases between states cannot be excluded. As per the census of India (2011), 29·9% of total human population are migrants and 13·8% of the total population migrates between states, possibly due to social, economic and political reasons^29^. Many anecdotal reports reveal interstate movement of many of these migrants due to national lockdown/curfew and loss of jobs; however, exact figures remain unknown. Interstate migration of COVID-19 positive cases resulted in an unexplained bias in state-level estimates. For example, Tamil Nadu state reported 646 cases as of 26 May 2020, with 54 cases being persons returning from other states^30^. Although these cases were not included for Tamil Nadu estimates, such data were not available from other states. Therefore, under or overestimation at the state level cannot be ruled out.

Many other epidemiologic variables such as symptom-onset date, proportion of asymptomatic or undiagnosed cases, as well as diagnostic testing patterns, remain unavailable. Moreover, impacts of individual state-level or component parts of the overall strategies during the government-imposed interventions remain unexamined. Therefore, current estimates should be carefully interpreted. Notwithstanding, in our opinion the current study provides much-needed information for further control and prevention of COVID-19 in India. Availability of additional data, hopefully in the near future, are expected to further improve these efforts.

## CONCLUSIONS

The Indian government imposed strict contact mitigation followed by phased relaxation, which slowed the spread of COVID-19 epidemic progression in India. These findings will also inform policy development for the control of COVID-19 epidemic in other regions and countries.

## Data Availability

All data referred to in the manuscript is available with the corresponding author of this manuscript.

## Contributions

BBS compiled and analysed the study data and prepared the first draft of the manuscript. ML, RTL, IAV, RD, JPSG, BSG and HWB supervised this study, refined the study objectives, corrected and interpreted the analysis, and revised the final manuscript.

## Acknowledgements

The authors acknowledge India’s National and State Health departments for collecting the daily COVID-19 epidemic data and releasing it in the public domain.

